# Poor Sleep Health Early After Stroke is Associated with Lower Quality of Life Later After Stroke

**DOI:** 10.1101/2025.05.30.25328681

**Authors:** George Fulk, Keenan Batts, Karen Klingman, Emily Peterson

## Abstract

**Background:** Factors early after stroke that are predictive of outcomes can help guide rehabilitation interventions. There is a growing understanding of the importance of sleep for recovery after stroke.

**Objectives:** To explore factors at 10 days post stroke, including sleep health (SH), that may be associated with QOL at 60 days post stroke.

**Methods:** Data from 62 participants were collected at 10 and 60 days post stroke. Independent variables at 10 days post stroke were the NIHSS, Barthel Index (BI), Montreal Cognitive Assessment (MOCA), Patient Health Questionnaire (PHQ-9), gait speed, Berg Balance Scale, and SH. SH was measured by combining data from actigraphy and self-report to create a score that reflected regularity, satisfaction, alertness, timing, efficiency, and duration of sleep (S-Ru-SATED). The Stroke Impact Scale (SIS) at 60 days post stroke assessed quality of life. A multiple linear regression using a leave-one-out cross validation was employed to assess the association between the independent variables at 10 days and the SIS at 60 days.

**Results:** The leave-one-out cross-validation analysis revealed that MOCA (p=0.0068), NIHSS (p=0.0181), BI (p=0.0028), and S-Ru-SATED (p=0.000155) at 10 days post stroke were significantly associated with the SIS at 60 days post stroke.

**Conclusions:** Participants with less stroke severity, higher cognition, better functional ability, and better SH early after stroke were more likely to have a higher QOL at 2 months post stroke. Research is needed to assess the impact of interventions to improve SH in conjunction with rehabilitation interventions after stroke.

## Introduction

Early discharge from the hospital and shortened length of stays for people who have experienced a stroke creates the need for accurate prognostic models of long-term outcomes to assist with developing an appropriate plan of care, discharge planning, patient/family education, and goal setting to optimize care after stroke. Knowledge related to factors in the first days to weeks after stroke that may influence recovery of the ability to perform activities of daily living (ADLs), walking ability, and quality of life (QOL) can be used not only to guide selection of optimal interventions and discharge planning, but also to optimize QOL for people recovering from stroke. A variety of studies have explored factors early after stroke that predict QOL, ADL performance, and walking ability. In a meta-analysis, Silva and colleagues^1^ found that ADL function early after stroke was a significant predictor of QOL. Initial severity of neurological deficits, upper extremity paresis, performance of ADLs, and age are predictive of ADL ability at three months post stroke.^2,3^ Lower extremity paresis and strength, trunk control, intact corticospinal tract, ADL ability, age, and balance early after stroke are predictive of independent walking ability later after stroke.^4,5^

Recent evidence points to the importance of sleep during recovery after stroke. People with concomitant sleep disorders, particularly obstructive sleep apnea (OSA), are more likely to exhibit greater disability than those without a sleep disorder.^6^ Fulk and colleagues^7^ found that people with stroke who self-reported sleep difficulties that interfered with their function had lower scores on the memory, emotion, communication, ADL, mobility, and participation subscales of the Stroke Impact Scale (SIS) than those that reported no problems with their sleep. Kasai and colleagues^8^ found that participants with self-reported poor quality of sleep after stroke had lower physical components of quality of life compared to participants that reported moderate to good quality of sleep. Kim and colleagues^9^ found that sleep duration was associated with QOL after stroke.

Given the impact of sleep health^10^ on overall health and its association with recovery after stroke, sleep early after stroke may influence future recovery. Previous research has not included sleep early after stroke when examining factors that may be associated with QOL later in recovery. The purpose of this study was to explore factors at 10 days post stroke, including sleep health, that may be associated with QOL at 60 days post stroke. We hypothesized that ADL function, stroke severity, depression, cognition, sleep health, balance, and walking ability at 10 days post stroke would be associated with QOL measured by the SIS at 60 days post stroke.

## METHODS

Participants were recruited from three inpatient rehabilitation hospitals, one in the Northeast, one in the Midwest, and one in the Southeast of the United States, and are part of a larger, ongoing cohort study to determine the prevalence of non-obstructive sleep apnea (OSA) sleep disorders and their impact on recovery after stroke.^11^ Inclusion criteria were diagnosis of stroke, >= 18 years old, National Institutes of Health Stroke Scale (NIHSS) item 1a score < 2 (alert or not alert but arousable by minor stimulation to obey, answer, or respond), provide informed consent or assent, with the participant’s legal guardian providing consent. Exclusion criteria were pre-stroke diagnosis of obstructive sleep apnea (OSA), oxygen desaturation index (ODI) >= 15 (ODI >=15 is an indication of possible moderate to severe OSA), living in nursing home or assisted living prior to stroke, unable to ambulate at least 150’ independently prior to stroke, other neurologic health condition that could impact recovery, pregnant, recent hemicraniectomy or suboccipital craniectomy, planned discharge > 150 miles from recruiting hospital, and global aphasia as determined by a score of 3 on NIHSS item 9. All participants provided verbal consent or assent (with participant’s legal guardian providing consent), and the study was approved by all local IRBs and a central IRB.

### Independent variables

At 10 days post stroke, while participants were undergoing inpatient rehabilitation, the National Institutes of Health Stroke Scale (NIHSS), Barthel Index (BI), Montreal Cognitive Assessment (MOCA), Patient Health Questionnaire (PHQ-9), gait speed (GS) measured at a comfortable pace over the middle 6 meters of a 10-meter walk, Berg Balance Scale (BBS), and sleep health, based on self-report measures and actigraphy (see below), were collected. Previous research has demonstrated that all these measures early after stroke are associated with QOL, ADL, and walking ability later in recovery. The NIHSS is a commonly used measure of stroke severity and previous research has shown that greater stroke severity early after stroke is associated with poorer ADL ability at three months post stroke.^2^ The BI measures basic mobility and ADL function. Previous research has shown that poorer ADL and mobility function early after stroke are associated with poorer QOL^1^ and limited walking ability^5^ later after stroke. The MOCA is a measure of cognitive ability and people with cognitive impairment in the first weeks after stroke are more likely to have greater disability and self-reported physical function at three months and one year after stroke.^12^ PHQ-9 is a self-report measure of depression. People with stroke who report greater symptoms of depression at two weeks post stroke are more likely to have poor QOL and greater disability at three and 12 months after stroke.^13^ Gait speed is associated with community walking activity.^14,15^ The BBS is a measure of functional balance and has been shown to predict independent walking after stroke.^4^

To measure sleep parameters that were used to measure sleep health, participants wore an ActiGraph wGT3X-BT (AG) (https://actigraphcorp.com/actigraph-wgt3x-bt/) 24 hours a day for 5 days on their least affected wrist to estimate sleep parameters.^16^ The AG was set to collect data at a 30Hz sample rate and later uploaded for analysis as 60 second epochs with three-axis motion detection. At the conclusion of each timepoint’s wear period, data were transferred from the AG to a study computer for analysis using ActiGraph’s ActiLife v6.13.4 software. Wear times were validated (marked as “wear” if either Troiano method or ActiGraph sensor indicated “wear”), and sleep parameters were determined via the Cole Kripke algorithm and Actigraph sleep period detection method. First-pass software-calculations of time-to-bed and time-out-of-bed were sometimes adjusted prior to calculation of sleep parameter variables, under certain conditions.

For overnight timeframes with two separate sleep periods detected by the software, a single sleep period spanning the combination of both originally detected periods was substituted. If the ActiLife software was unable to determine time to bed and time out of bed, every effort was made to manually determine these from activity and light signals in the ActiLife interface. Any sleep periods appearing to be daytime naps were not included in analysis of nightly sleep. The following sleep parameters were derived from the AG: time to bed, time awakened, total sleep time, sleep efficiency, wake after sleep onset (WASO), and number of awakenings (NOA).

Participants also completed two sleep related questionnaires, the Epworth Sleepiness Scale (ESS)^17,18^ and the Insomnia Severity Index (ISI).^19,20^

To measure sleep health, we combined sleep parameter data derived from the AG and self-report data from the ESS and ISI to create a 12-point Surrogate Ru-SATED (S-Ru-SATED) score adapted from the framework of sleep health proposed by Buysse.^10^ The sleep parameters components of the S-Ru-SATED from the AG were Regularity, Timing, Efficiency, and Duration. For Satisfaction we used the score on question on the ISI, which asks participants to rate how satisfied/dissatisfied they are with their current sleep pattern. Alertness was determined by the ESS score. Sleep regularity was scored based on consistency in bedtimes and wake times, with a score of 2 assigned if deviations were within ±1 hour of the median of bedtime and wake time every night and morning, 1 if this occurred on all but one night or morning, and 0 for two or more deviations. Timing was scored by whether participants slept between 2–4 am, with scores of 2, 1, and 0 assigned for meeting this criterion every night, all but one night, and more than one night, respectively. Efficiency was scored based on nightly sleep efficiency, with 2 assigned if all nights were ≥85%, 1 for all but one night, and 0 for two or more nights <85%. Duration score was based on total sleep time, with a score of 2 for sleeping 7–9 hours each night, 1 for all but one night, and 0 for two or more nights outside this range. Satisfaction was determined from the ISI question about sleep satisfaction, with scores of 2, 1, and 0 corresponding to “very satisfied” or “satisfied,” “somewhat dissatisfied,” and “dissatisfied” or “very dissatisfied,” respectively. Alertness was scored using the ESS, with scores of 2 for ESS <11, 1 for scores of 11–12, and 0 for scores >12. The total S-Ru-SATED score ranged from 0 to 12, with higher scores indicating better sleep health, table 1.

**Table 1.** Surrogate Ru-SATED Scale.

| **Item and Definition from Ru-SATED<sup>a</sup>** | **S-Ru-SATED<sup>b</sup>** |  |  | **Ru-SATED<sup>a</sup>** |  |  |
| --- | --- | --- | --- | --- | --- | --- |
| **Regularity:** | 2: Participant went to bed and got out of bed every night and morning within 1 hour of the median time went to bed and got out of bed. | 1: Participant went to bed and got out of bed all but one night or morning within 1 hour of the median time went to bed and bed and got out of bed. | 0: Participant went to bed and got out of bed all more than one night or morning within 1 hour of the median time went to bed and bed and got out of bed. | 2: Usually/Always | 1: Sometimes | 0: Rarely/Never |
| **Satisfaction:**<br>Are you satisfied with your sleep? | 2: 0 or 1 ISI<sup>c</sup><br>satisfaction with sleep question | 1: 2 on ISI<sup>c</sup><br>satisfaction with sleep question | 0: 3 or 4 on ISI<sup>c</sup><br>satisfaction with sleep question | 2: Usually/Always | 1: Sometimes | 0: Rarely/Never |
| **Alertness:**<br>Do you stay awake all day without dozing? | 2: <11 on ESS<sup>d</sup> | 1: 11 or 12 on the ESS<sup>d</sup> | 0: >12 on the ESS<sup>d</sup> | 2: Usually/Always | 1: Sometimes | 0: Rarely/Never |
| **Timing:** Are you asleep (or trying to sleep) between 2a and 4a? | 2: Participant was in bed every night between 2a and 4a. | 1: Participant was in bed all but one night between 2a and 4a. | 0: Participant was not in bed more than one night between 2a and 4a. | 2: Usually/Always | 1: Sometimes | 0: Rarely/Never |
| **Efficiency:**<br>Do you spend less than 30 minutes awake at night? | 2: Sleep efficiency 85% or greater every night. | 1: Sleep efficiency 85% or greater all but one night. | 0: Sleep efficiency less than 85% two or more nights. | 2: Usually/Always | 1: Sometimes | 0: Rarely/Never |
| **Duration:**<br>Do you sleep between 6 and 8 hours per day? | 2: Total sleep time between 7 and 9 hours each night. | 1: Total sleep time between 7 and 9 hours for all but one night. | 2: More than one night during which total sleep time was not between 7 | 2: Usually/Always | 1: Sometimes | 0: Rarely/Never |
|  |  |  | and 9 hours. |  |  |  |

<sup>a</sup>Ru-SATED: Regularity, Satisfaction, Timing, Efficiency, Duration; <sup>b</sup>S-Ru-SATED: Surrogate, Regularity, Satisfaction, Timing, Efficiency, Duration; <sup>c</sup>Insomnia Severity Index; <sup>d</sup>Epworth Sleepiness Scale.

Additionally, the following data were collected at 10 days post stroke: age, sex, marital status, race, time since stroke, type of stroke, and location of stroke to describe the sample.

### Dependent variable

The SIS was administered through an in person interview at 60 days post stroke. The SIS is a self-report questionnaire that measures QOL in individuals after stroke.^21–23^ It contains 9 subscales and a single item that asks participants to rate the percentage of their overall recovery compared to prior to their stroke. We used the total SIS score in these analyses.

### Data Analysis

Descriptive statistics were used to describe the sample (age, sex, marital status, race, time since stroke, type of stroke, location of stroke, length of stay in rehabilitation, and if and how much physical, occupational, and speech therapy participants received between discharge from the hospital and 60 days post stroke).

A multiple linear regression using a leave-one-out cross validation was employed to assess the association between the independent variables described above (NIHSS, BI, MOCA, PHQ-9, GS, BBS, S-Ru-SATED, NOA, and WASO) taken at 10 days post stroke and SIS measured at 60 days post stroke. This technique involves iteratively using each data point in the dataset as a single test case while the remaining data points form the training set. Specifically, for a dataset with n observations, the model is trained n times, each time leaving out one observation for testing and using the remaining n − 1 observations for training. The process is repeated until each observation has been used exactly once as the test set. R version 4.4.2 was used for all analyses and the *caret* package was used for the leave-one-out cross validation.

## Results

The analyses used data from 62 participants. The mean (standard deviation) age was 64.6 (16.8) years and 55% of the participants were females. Independent variables were collected at a mean of 9.4 (7.1) days post stroke, see Table 2. The mean (standard deviation) of the SIS collected at a mean of 60.0 (4.0) days after stroke was 0.73 (0.16), see Table 3.

**Table 2.** Demographic Information at 10 Days Post Stroke.

| Characteristic | N = 62 |
| --- | --- |
| Sex |  |
| Male | 28 / 62<br>(45%) |
| Female | 34 / 62<br>(55%) |
| Age | 64.57<br>(16.80) |
| Race |  |
| Black | 11 / 62<br>(18%) |
| White | 50 / 62<br>(81%) |
| More than one race | 1 / 62<br>(1.6%) |
| Ethnicity |  |
| Hispanic or Latino | 1 / 62<br>(1.6%) |
| Not Hispanic or Latino | 61 / 62<br>(98%) |
| Stroke Type |  |
| Ischemic | 49 / 62<br>(79%) |
| Hemorrhagic | 12 / 62<br>(19%) |
| Unknown | 1 / 62<br>(1.6%) |
| Location of Stroke |  |
| Left Hemisphere | 3 / 62<br>(4.8%) |
| Right Hemisphere | 18 / 62<br>(29%) |
| Left Subcortical | 8 / 62 |
|  | (13%) |
| Right Subcortical | 10 / 62<br>(16%) |
| Brainstem | 15 / 62<br>(24%) |
| Cerebellar | 7 / 62<br>(11%) |
| Bilateral | 1 / 62<br>(1.6%) |
| Days from stroke to admission to inpatient rehabilitation hospital | 9.3 (7.1) |
| Days from stroke to discharge from inpatient rehabilitation hospital | 28.9 (11.6) |
| Length of Stay (days) | 19.5 (8.9) |
| Berg Balance Scale | 14.2 (14.2) |
| Barthel Index | 39.1 (21.5) |
| Gait speed, meters/second | 0.21 (0.29) |

| **Characteristic** | **N = 62** |
| --- | --- |
| Montreal Cognitive Assessment | 21.3 (5.4) |
| National Institutes of Health Stroke Scale | 5.3 (3.6) |
| Patient Health Questionnaire | 3.9 (4.3) |
| Surrogate Ru-SATED | 5.0 (2.1) |
| Number of Awakenings/nights | 13.3 (6.0) |
| Wake After Sleep Onset (minutes) | 82.4 (49.5) |

**Table 3.** Demographic Information at 60 Days Post Stroke.

| **Characteristic** | **60 days**<br>N = 62 |
| --- | --- |
| Living Situation |  |
| Home with family/caregiver | 55 / 62 (89%) |
| Home alone | 4 / 62 (6.4%) |
| Subacute rehab | 3 / 62 (4.7%) |
| Days from stroke to 60 days follow up, mean (sd) | 60 (4) |
| Receiving rehabilitation services | 56 / 62 (90%) |
| Stroke Impact Scale, mean (sd) | 73.0 (16.0) |

The leave-one-out cross-validation analysis revealed that four variables, MOCA (p=0.0068), NIHSS (p=0.0181), BI (p=0.0028), and S-Ru-SATED (p=0.000155) at 10 days post stroke were significantly associated with the SIS at 60 days post stroke. The model demonstrated a strong fit, with an adjusted R-squared value of 0.6618, indicating that approximately 66.18% of the variance in the SIS could be explained by the independent variables. The model’s predictive accuracy was further supported by the root mean square error (RMSE) and mean absolute error (MAE) values. The RMSE was 0.6068, suggesting that the average deviation of the predicted values from the observed values was relatively low. Additionally, the MAE was 0.0784, indicating a high level of precision in the model’s associations.

## DISCUSSION

The present study explored which measures early after stroke were associated with QOL at two months post stroke. We found that the MOCA, NIHSS, BI, and S-Ru-SATED at 10 days post stroke were significantly associated with the QOL as measured by the SIS at 60 days post stroke. These factors explained 66% of the variance in the SIS. Individuals with less stroke severity, higher cognition, better functional ability, and better sleep health early after stroke were more likely to have a higher QOL at 2 months post stroke.

Our findings that cognition,^24^ functional ability,^1^ and stroke severity^25,26^ early after stroke are associated with QOL later in recovery are similar to what others have reported. Although we hypothesized that walking ability and balance would also be associated with quality of life this did not bear out in our results. Walking ability and functional balance are critical components of overall functional ability and could have been accounted for by the BI. Our finding that SH early after stroke was also associated with quality of life later in recovery is novel. To the best of our knowledge this has not been explored previously. Sleep is critical for health and there is a growing body of evidence of its importance in people who have experienced a stroke. People with the sleep disorders and those that report poor quality sleep are likely to have poorer outcomes compared to those without sleep disturbances.^6,7^ Our findings provide further support of the importance of healthy sleep during recovery after stroke.

Most of the research related to the impact of sleep in people with stroke has focused on specific sleep disorders and less attention has been paid to general sleep health. It may be important to examine sleep from this broader view as well, particularly during the early stages of recovery. Nonpharmacological interventions designed to improve sleep quality while in the hospital such sleep hygiene, sleep aids, relaxation techniques, and music have shown benefits in the people who are hospitalized for a variety of health conditions.^27,28^ Extending these interventions and combining them with active rehabilitation in people with stroke may help improve recovery.

A limitation of our study is that people with stroke who have moderate to severe OSA were not included in this study. The results need to be interpreted with this in mind. Obstructive sleep apnea is common in people with stroke and people with OSA likely have poor SH.

### Conclusion

Sleep health, cognition, stroke severity, and functional ability at 10 days post stroke were associated with self-reported quality of life at 60 days post stroke. These factors explained 66% of the variance in quality of life. Rehabilitation professionals should consider incorporating interventions to improve sleep health in conjunction with active rehabilitation interventions during recovery after stroke.

## Data Availability

Data produced for this study are not currently available.

## Notes

**Funding Support:** This work was supported by the National Institutes of Health NINR, Award Number R01NR018979

### Competing Interest Statement

The authors have declared no competing interest.

### Funding Statement

This work was funded by the National Institutes of Health NINR, Award Number R01NR018979

### Author Declarations

This research was approved by the following Institutional Review Boards: WCG IRB, Upstate Medical University IRB, University of Kansas Medical Center IRB, and Emory University IRB.

## References

1. Silva AC, Menezes KKP, Scianni AA, Avelino PR, Faria C. Predictors of health-related quality of life one year after stroke: a systematic review with meta-analysis. Int J Rehabil Res. Jun 1 2024;47(2):53–63. doi:10.1097/MRR.0000000000000623

2. Veerbeek JM, Kwakkel G, van Wegen EE, Ket JC, Heymans MW. Early prediction of outcome of activities of daily living after stroke: a systematic review. Stroke. May 2011;42(5):1482–8. doi:10.1161/STROKEAHA.110.604090

3. Heuschmann PU, Wiedmann S, Wellwood I, et al. Three-month stroke outcome: the European Registers of Stroke (EROS) investigators. Neurology. Jan 11 2011;76(2):159–65. doi:10.1212/WNL.0b013e318206ca1e

4. Wouda NC, Knijff B, Punt M, Visser-Meily JMA, Pisters MF. Predicting Recovery of Independent Walking After Stroke: A Systematic Review. Am J Phys Med Rehabil. May 1 2024;103(5):458–464. doi:10.1097/PHM.0000000000002436

5. Preston E, Ada L, Stanton R, Mahendran N, Dean CM. Prediction of Independent Walking in People Who Are Nonambulatory Early After Stroke: A Systematic Review. Stroke. Oct 2021;52(10):3217–3224. doi:10.1161/STROKEAHA.120.032345

6. Fulk GD, Boyne P, Hauger M, et al. The Impact of Sleep Disorders on Functional Recovery and Participation Following Stroke: A Systematic Review and Meta-Analysis. Neurorehabil Neural Repair. Nov 2020;34(11):1050–1061. doi:10.1177/1545968320962501

7. Fulk G, Duncan P, Klingman KJ. Sleep problems worsen health-related quality of life and participation during the first 12 months of stroke rehabilitation. Clin Rehabil. Nov 2020;34(11):1400–1408. doi:10.1177/0269215520935940

8. Kasai A, Saitou H, Takano M, et al. Pre-stroke habitual prolonged sleep as a predictor for post-stroke sleep quality, stroke-related quality of life, and lifestyle values. J Clin Neurosci. Aug 2021;90:26–31. doi:10.1016/j.jocn.2021.05.018

9. Kim Y, Moon HM. Association between quality of life and sleep time among community-dwelling stroke survivors: Findings from a nationally representative survey. Geriatr Gerontol Int. Dec 2019;19(12):1226–1230. doi:10.1111/ggi.13797

10. Buysse DJ. Sleep health: can we define it? Does it matter? Sleep. Jan 1 2014;37(1):9–17. doi:10.5665/sleep.3298

11. Klingman KJ, Skufca JD, Duncan PW, Wang D, Fulk GD. Study Protocol: Sleep Effects on Poststroke Rehabilitation Study. Nurs Res. Nov-Dec 01 2022;71(6):483–490. doi:10.1097/nnr.0000000000000611

12. Liao X, Zuo L, Dong Y, et al. Persisting cognitive impairment predicts functional dependence at 1 year after stroke and transient ischemic attack: a longitudinal, cohort study. BMC Geriatr. Dec 31 2022;22(1):1009. doi:10.1186/s12877-022-03609-z

13. Wulsin L, Alwell K, Moomaw CJ, et al. Comparison of two depression measures for predicting stroke outcomes. J Psychosom Res. Mar 2012;72(3):175–9. doi:10.1016/j.jpsychores.2011.11.015

14. Fulk GD, He Y, Boyne P, Dunning K. Predicting Home and Community Walking Activity Poststroke. Stroke. Feb 2017;48(2):406–411. doi:10.1161/STROKEAHA.116.015309

15. Bijleveld-Uitman M, van de Port I, Kwakkel G. Is gait speed or walking distance a better predictor for community walking after stroke? J Rehabil Med. Jun 2013;45(6):535–40. doi:10.2340/16501977-1147

16. Smith MT, McCrae CS, Cheung J, et al. Use of Actigraphy for the Evaluation of Sleep Disorders and Circadian Rhythm Sleep-Wake Disorders: An American Academy of Sleep Medicine Clinical Practice Guideline. J Clin Sleep Med. Jul 15 2018;14(7):1231–1237. doi:10.5664/jcsm.7230

17. Mills RJ, Koufali M, Sharma A, Tennant A, Young CA. Is the Epworth sleepiness scale suitable for use in stroke? Top Stroke Rehabil. Nov-Dec 2013;20(6):493–9. doi:10.1310/tsr2006-493

18. Johns MW. A new method for measuring daytime sleepiness: the Epworth sleepiness scale. Sleep. Dec 1991;14(6):540–5.

19. Morin CM, Belleville G, Belanger L, Ivers H. The Insomnia Severity Index: psychometric indicators to detect insomnia cases and evaluate treatment response. Sleep. May 1 2011;34(5):601–8. doi:10.1093/sleep/34.5.601

20. Bastien CH, Vallieres A, Morin CM. Validation of the Insomnia Severity Index as an outcome measure for insomnia research. Sleep Med. Jul 2001;2(4):297–307. doi:10.1016/s1389-9457(00)00065-4

21. Lai SM, Perera S, Duncan PW, Bode R. Physical and social functioning after stroke: comparison of the Stroke Impact Scale and Short Form-36. Stroke. Feb 2003;34(2):488–93.

22. Duncan PW, Bode RK, Min Lai S, Perera S. Rasch analysis of a new stroke-specific outcome scale: the Stroke Impact Scale. Arch Phys Med Rehabil. Jul 2003;84(7):950–63. doi:S0003999303000352 [pii]

23. Duncan PW, Wallace D, Lai SM, Johnson D, Embretson S, Laster LJ. The stroke impact scale version 2.0. Evaluation of reliability, validity, and sensitivity to change. Stroke. Oct 1999;30(10):2131–40.

24. Zietemann V, Georgakis MK, Dondaine T, et al. Early MoCA predicts long-term cognitive and functional outcome and mortality after stroke. Neurology. Nov 13 2018;91(20):e1838–e1850. doi:10.1212/WNL.0000000000006506

25. Yeoh YS, Koh GC, Tan CS, et al. Can acute clinical outcomes predict health-related quality of life after stroke: a one-year prospective study of stroke survivors. Health Qual Life Outcomes. Nov 21 2018;16(1):221. doi:10.1186/s12955-018-1043-3

26. Chen CM, Tsai CC, Chung CY, Chen CL, Wu KP, Chen HC. Potential predictors for health-related quality of life in stroke patients undergoing inpatient rehabilitation. Health Qual Life Outcomes. Aug 5 2015;13:118. doi:10.1186/s12955-015-0314-5

27. Herscher M, Mikhaylov D, Barazani S, et al. A Sleep Hygiene Intervention to Improve Sleep Quality for Hospitalized Patients. Jt Comm J Qual Patient Saf. Jun 2021;47(6):343–346. doi:10.1016/j.jcjq.2021.02.003

28. Beswick AD, Wylde V, Bertram W, Whale K. The effectiveness of non-pharmacological sleep interventions for improving inpatient sleep in hospital: A systematic review and meta-analysis. Sleep Med. Jul 2023;107:243–267. doi:10.1016/j.sleep.2023.05.004

